# Psychometric Properties and Longitudinal Measurement Invariance of the Spanish Version of the Alcohol Expectancies Questionnaire – Short Form among Young Adult Binge Drinkers

**DOI:** 10.1101/2023.11.14.23298472

**Authors:** Bella M. González-Ponce, Angelina Pilatti, Gabriela Rivarola Montejano, Adrian J. Bravo, Fermín Fernández-Calderón

**Author notes:** Corresponging author: Bella María González Ponce. Address: Department of Clinical and Experimental Psychology, University of Huelva. Office number PB 3-12. Campus de «El Carmen». Avenida de las Fuerzas Armadas, S/N. 21071, Huelva, Spain. **Disclosure statement:** The authors report there are no competing interests to declare.

## Abstract

**Background:** Longitudinal Measurement Invariance (LMI) is critically important to evaluate changes in the alcohol expectancies over time. However, few studies have yet explored the longitudinal properties of the Spanish EQ-SF.

**Objectives:** To examine the reliability, sources of validity (structural and concurrent validity), and LMI of the Spanish short version of the Alcohol Expectancy Questionnaire in a sample of young adults who engage in binge drinking.

**Methods:** The participants (*n* = 279; Mean age = 21.33, *SD* = 2.15; 48.4% female) completed the Spanish EQ-SF, and two months later they completed this measure again, along with measures to alcohol use, drinking motives, and protective behavioral strategies (PBS). Confirmatory Factor Analysis was used to identify which of two proposed models provided the best-fitting factor structure. We aimed to determine whether the best-fitting model was invariant across assessments and to evaluate the predictive validity and reliability of the scores.

**Results:** Our findings revealed that the eight-factor intercorrelated model provided the best fit. This model was invariant across assessments, providing evidence for longitudinal measurement invariance. Moreover, the scores showed adequate reliability (.68 to .90) and predictive validity (i.e., positive alcohol expectancies were positively related to alcohol use and drinking motives and negatively related to PBS).

**Conclusion:** Our results support the reliability, validity, and temporal invariance of the EQ-SF scores among Spanish young adults with binge drinking patterns. The evidence supports the suitability of this measure for accurately assessing changes in alcohol expectancies over time in interventions aimed at preventing binge drinking in young adults.

## Introduction

Heavy drinking is positively associated with various alcohol-related negative consequences (1). Binge drinking, which is typically defined as the consumption of 4/5 standards drinks (women/men) within ≤ 2 hours (2), is a prevalent behavior among emerging adults worldwide (3). To illustrate, a study with college students from the U.S., Spain, and Argentina who reported alcohol use within the previous month revealed that, overall, students reported consuming alcohol drink 6 days per month, with 2 of these occasions involving binge drinking episodes (4). In Spain, alcohol ranks as the most frequently used psychoactive substance among individuals aged between 15 and 65. Among young adults (i.e., 18-25 years) binge drinking is highly prevalent, reaching 35.4% among men and 24.1% among women compared to 15.4% in the overall population (5).

Engaging in binge drinking can be, at least in part, explained and predicted by alcohol expectancies. Rooted in the framework of Social Learning Theory (6), alcohol expectancies refer to learned associations about the effects of alcohol consumption on behavior, mood, and emotions (7). Individuals, either through their own experience with alcohol or by observing others (8), develop “if … then” expectations (e.g., *if I drink alcohol then I would feel less nervous at a party*) that influence alcohol consumption.

Alcohol expectancies are considered proximal predictors with a direct impact on alcohol use, including binge drinking (9), that also mediate the indirect effect of individual and environmental predictors (10). For instance, alcohol expectancies have been identified as significant mediators in the relationship between diverse distal factors such as an earlier age of first intoxication (11), personality traits (12), social norms (13), and greater alcohol use and related problems.

Alcohol expectancies are typically categorized into two broad groups: positive and negative (14). Positive alcohol expectancies encompass beliefs about the rewarding and desirable effects of alcohol use (e.g., social facilitation, feeling calm and relaxed, or feeling more sexually appealing) and have been positively associated with drinking initiation (15), higher levels of alcohol consumption (16), greater occurrence of binge drinking (9), and more alcohol-related negative consequences (17). Moreover, positive alcohol expectancies are negatively associated with the use of Protective Behavioral Strategies ([PBS], specific behaviors implemented to mitigate the harmful consequences of alcohol consumption [18]), ultimately leading to more alcohol-related negative consequences (19). In contrast, negative alcohol expectancies, which include beliefs about physical, cognitive, and emotional impairment (e.g., feeling guilty, sick, or confused), have been associated with limited alcohol use or complete abstinence (20).

A key feature of alcohol expectancies is their capacity for change, and they have been targeted in interventions aimed at reducing or preventing alcohol use by manipulating (i.e., changing) these beliefs (21). Given the relevance of alcohol expectancies in understanding drinking involvement, coupled with the potential to reduce alcohol-related problems by targeting these beliefs, this construct has attracted increasing interest among the research community. Thus, a substantial body of research has been dedicated to accurately assessing alcohol expectancies. While various measurement tools have been developed (e.g., the Alcohol Expectancy Questionnaire [AEQ] [22], the Comprehensive Effects of Alcohol [CEOA] [23]), the Expectancy Questionnaire (EQ) (14) offers several notable advantages. Unlike the AEQ, the EQ assesses both positive and negative alcohol expectancies, and, in contrast to the CEOA, it includes two separate dimensions for assessing both cognitive and physical impairment. This approach better discriminates between these two forms of harm, which have been differentially associated to alcohol use (24).

Additionally, and relevant to the present study, a Spanish adaptation of this measure was validated in a sample of Spanish adolescents, with the findings providing supporting evidence of validity and reliability of the measure (24). Despite these advantages, this measure could be lengthy to administer, particularly in research or clinical settings where individuals are repeatedly asked to answer the same questions. This feature could potentially deter individuals from completing the assessment. To address this concern, Mezquita et al. (25) developed and validated a brief version of the Spanish EQ in two separate samples of adolescents and adults (EQ-SF). By employing a combination of Classical Test Theory and Item Response Theory analyses, these authors obtained a reduced version comprising of 24 items loaded onto the eight original dimensions of the EQ, grouped into two second-order factors: positive expectancies (social positive, fun, sex, and tension reduction) and negative expectancies (social negative, emotional negative, physical negative, and cognitive negative).

Different sources of validity, adequate reliability, and measurement invariance across sex (male, female) and age (adolescents, adults) indicated the EQ-SF is an adequate measure for evaluating alcohol expectancies in Spanish youths and adults.

### Purpose of Present Study

When validating a scale, particularly in the context of repeated assessments, it is important to examine the stability of scores over time, a concept known as Longitudinal Measurement Invariance (LMI). LMI serves as a method to determine if the instrument consistently captures the underlying construct across different time points (26). A consistent LMI suggests that the assessment remains relatively stable over time (27) so that any changes in mean scores can be taken to indicate genuine changes in the construct levels (28). In general, tests of LMI involve three sequential steps, each introducing additional equality constraints on the parameters (29). These levels of invariance are configural (i.e., whether the factor structure shows adequate fit across time points), metric (i.e., if factor loadings are equivalent over time), and scalar (whether the item intercepts remain equivalent across time points) (30). As indicated previously, research has established the measurement invariance of the Spanish EQ-SF across sex and age (25). However, limited research has investigated the invariance of the EQ-SF across time and in a high-risk population (i.e., emerging adults who engage in binge drinking). Therefore, the present study aimed to examine the LMI properties of the EQ-SF in a sample of young adults who have reported engaging in binge drinking in the past two months. Specifically, we aimed to examine LMI and test two models with different factorial structures: one with eight correlated factors and another where these subscales were grouped into two (positive and negative) second-order factors.

Additionally, we conducted reliability estimations of the scale scores at both baseline (Time 1) and follow-up (Time 2) in our sample. To provide evidence of the scale’s validity based on its relationship with other variables, we analyzed the EQ-SF scores at Time 1 with respect to alcohol-related outcomes (e.g., frequency of alcohol use and alcohol-related consequences), drinking motives, and the use of PBS at Time 2.

## Method

### Participants and Procedure

Participants were 360 young adults (*Mean age* = 21.1 years; *SD* = 2.21) who were part of a longitudinal research project (31) approved by the Regional Bioethics Research Committee of Andalusia (Consejería de Sanidad, Government of Andalusia, Spain). Participants were selected through a targeted sampling procedure from various community settings, including parks, pubs, bars, and discotheques in the province of Huelva (Spain). The inclusion criteria were as follows: a) being between 18 and 25 years old, b) a history of alcohol use on two or more occasions within the past month, and c) agreeing to participate in the two-month follow-up. A psychosocial psychologist recruited participants who met these criteria from the selected settings, and information about the study was also disseminated through posters displayed within those settings. The targeted sampling procedure (32) often utilizes the participants’ social networks to identify new candidates (a form of snowball sampling, as described by Goodman [33]). Consequently, using this procedure, a total of 360 participants were recruited for the initial assessment, of which 174 (48.3%) were recruited directly by the interviewer in the selected contexts, 155 participants (43.1%) were nominated from the participants’ social network, and 31 (8.6%) contacted the interviewer after seeing the posters in the selected contexts.

Participants completed the questionnaires for the first session (Time 1 [T1]) in paper and pencil format at the University of Huelva. Before their participation, all participants gave their informed consent, and upon completing the questionnaires, they received compensation in the form of an Amazon voucher worth 15 euros. Those who completed the follow-up questionnaire (Time 2 [T2]) after two months also received a 15-euro Amazon voucher. Most of the sample (*n* = 339, 94.2%) participated in the two-month follow-up assessment.

For the purposes of the present study, we selected those participants who reported having consumed 5 or more alcoholic drinks (for males) or 4 or more alcoholic drinks (for females) within a 2-hour period on at least one occasion in the past two months. A subgroup of 279 (93.94%) young adults of the total sample engaged in binge drinking and returned to complete the EQ-SF and other related measures at the follow-up assessment, which took place two months after the initial assessment. Thus, the analytic sample comprised of 279 young adults who engaged in binge drinking (*Mean age* = 21.33, *SD* = 2.15; 51.6% male). The vast majority of participants (96.3%) reported being born in Spain and attending university (87.2%) at the time of participating in the study. Most of the participants (78.5%) lived with their parents, with the primary source of income being family allowance (51.9%), and 30.3% were studying and working. The mean number of binge drinking days at both the baseline and follow-up assessments was 6.9 (*SD* = 8.4) and 4.6 (*SD* = 5.7), respectively.

### Instruments

#### Alcohol expectancies (at baseline and follow-up)

The Spanish Short Form (EQ- SF) (25) of the Expectancies Questionnaire (14) comprises 24 items and employs a Likert response scale with 6 options, ranging from 0 (never) to 5 (always), to measure positive and negative expectancies. This scale is composed of 8 dimensions, 4 of which correspond to positive expectations (12 items): expectancies about social facilitation (Social Positive), positive affect potentiation (Fun Positive), sexual disinhibition (Sex Positive), and tension reduction (Tension Reduction). The remaining four dimensions correspond to negative expectations (also comprising 12 items): expectancies about the antisocial effects of alcohol (Social Negative), negative emotional states (Emotional Negative), as well as undesirable physical (Physical Negative) and cognitive effects (Cognitive Negative). Participants were asked to indicate the likelihood of experiencing the aforementioned consequences when consuming alcohol. The reliability of the Spanish version of the EQ-SF (25) was adequate, with Cronbach’s alpha values ranging between .77 and .93.

#### Alcohol consumption (at follow-up)

At follow-up, we collected information on frequency of drinking, and drunkenness and binge drinking episodes. For each of these frequency measures, items measured the number of days within the past two months during which participants had engaged in: alcohol consumption, drunkenness, and binge drinking. Binge drinking was defined as consuming ≥5drinks (in men) or ≥4drinks (in women) within a two-hour interval (2, 34).

#### Quantity of alcohol consumed in a typical week during the past month (at follow-up)

The modified version of the Daily Drinking Questionnaire (DDQ) (35) was used to assess the amount of alcohol consumed in a typical week during the past month. This questionnaire gathers information on the consumption of six types of alcoholic beverages, each accompanied by visual representations, according to the Spanish Observatory of Drugs and Addictions (36). The number of drinks consumed by participants was converted into Standard Drinking Units (SDUs). In Spain, 1 SDU equals 10 grams of pure alcohol (37).

#### Negative alcohol-related consequences (at follow-up)

We used the 48-item Spanish version (S-YAACQ) (38) of the Young Adult Alcohol Consequences Questionnaire (YAACQ) (39). Each item assesses the presence or absence (0 = no, 1 = yes) of negative alcohol-related consequences in the last month. The total score indicates the number of consequences experienced in that period. As recommended by the authors of the original scale (39), tetrachoric correlations were used to estimate internal consistency Ordinal alpha values were .95 (follow-up).

#### Drinking Motives (at follow-up)

We used the Spanish version (DMQ-R SF) (40) of the Drinking Motives Questionnaire-Revised (DMQ-R SF) (41). This instrument consists of 12 items grouped into four dimensions (with three items per dimension): social motives, coping motives, enhancement motives, and conformity motives. The response format was a 5-point scale (1 =L*almost never or never* to 5 =L*almost always or always*). Cronbach’s alpha values were as follows: social motives α = 0.82; enhancement motives α = 0.77; coping motives α = 0.80; conformity motives α = 0.83.

#### Protective Behavioral Strategies (at follow-up)

We used the Spanish version (S- PBSS-20) (42) of the Protective Behavioral Strategies Scale (PBSS-20) (43). This scale consists of 20 items grouped into three dimensions: Manner of Drinking (MOD-5 items), Stopping/Limiting Drinking (SLD-7 items), and Serious Harm Reduction (SHR- 8 items). Participants reported using PBS within the last two months using a Likert-type response format ranging from 1 (*never*) to 5 (*always*). Consistent with the Spanish version of the PBSS (42), internal consistency was estimated using McDonald ordinal alpha (MOD α = 0.65; SLD α = 0.69; SHR α = 0.69).

### Data analysis

A confirmatory factor analysis (CFA) was conducted to examine the internal structure of the EQ-SF, and two models with different factorial structures were evaluated. The first model (Model 1) comprises eight correlated factors, each with three items as observable variables. The second model (Model 2) is structured around two second-order factors (each with 12 items), with four dimensions per factor, and three items per dimension. These models were estimated using the Robust Maximum Likelihood Estimator (MLR) method. The following goodness-of-fit indicators were used to assess model fit: the χ2 statistic, the comparative fit index (CFI), and the Tucker-Lewis index (TLI), with values >.90 considered acceptable and >.95 as optimal. The root mean square error of approximation (RMSEA) and the standardized root mean square residual (SRMR) were also used, with values ≤ .10 indicating acceptable fit and values ≤ .08 indicating good fit (44, 45). Additionally, the fit of both models was compared by applying the Chi-Square Difference Testing using the Satorra-Bentler Scaled Chi-Square (46). To estimate the internal consistency of each subscale of the EQ-SF, Cronbach’s alpha coefficients were calculated separately for each subscale at the two time points.

The longitudinal invariance analysis of the EQ-SF was conducted across the two time points. Three levels of invariance were examined: configurational (i.e., whether the items load onto the proposed factors), metric (i.e., whether the factor loadings remain consistent across time), and scalar (to establish whether the factor loadings are stable, and the intercepts remain constant) (47). The CFI, TLI, and RMSEA indices were utilized to evaluate model fit, along with the previously mentioned reference values (44, 45). In addition, the change in model fit was examined from one model to another (ΔCFI and ΔTLI ≥ -.010, ΔRMSEA ≥ .015 and ΔSRMR ≥ .010) (48). To provide evidence of criterion-related validity, based on the relationship with other variables, Pearsońs correlations were employed to examine the between EQ-SF dimensions measured at T1 and five alcohol-related outcomes (frequency of alcohol use, frequency of drunkenness, frequency of binge drinking, quantity of alcohol consumed, and alcohol-related consequences), drinking motives (four dimensions: social, enhancement, coping, and conformity), and use of PBS (three dimensions: manner of drinking, stopping/limiting drinking, serious harm reduction) measured at T2. M*plus* 8.0 (49) was used for CFA and LMI analyses and SPSS 27 (51) was used to estimate Cronbach’s alpha values and conduct correlation analyses.

## Results

### Confirmatory factor analysis and reliability

Table 1 displays the fit measures of the two tested models. Both models showed an acceptable fit to the data according to most indices (CFI, TLI, and RMSEA). Notably, Model 1 (eight correlated factors) showed better fit in SRMR index (< .06) compared to Model 2 (two second-order factors; SRMR < .08). The Satorra-Bentler (46) chi-square difference test revealed that the model with eight correlated factors showed a significantly better fit than the second-order two-factor model (Δchi (Δdf) = 60.83 [19], ΔCFI = .00).

**Table 1.** Fit statistics: Confirmatory Factor Model of EQ-SF.

| Models | $\chi^2$ | *df* | *p* | CFI | TLI | RMSEA | SRMR |
| --- | --- | --- | --- | --- | --- | --- | --- |
| Model 1: Eight correlated factors | 395.59 | 224 | .000 | 0.929 | 0.913 | 0.051 | 0.057 |
| Model 2: Two second order factors | 456.32 | 243 | .000 | 0.912 | 0.900 | 0.054 | 0.072 |

*Note.* $\chi^2$ = chi-square; *df* = degree of freedom; *p* = p value; *CFI* = Comparative Fit Index; *TLI* = Tucker-Lewis Index; *SRMR* = Standardized Root Mean-Square Residual; *RMSEA* = Root Mean-Square Error of Approximation

Table 2 shows the factor loadings for the EQ-SF eight correlated factors model, organized according to items and time of assessment (T1 and T2). All item loadings were moderate to high across both assessment times, ranging from .50 to .92. The reliability coefficients, as measured by Cronbach’s alpha, for each dimension were consistently strong at both time points (from .69 to .90 at T1 and .68 to .89 at T2).

**Table 2.** Factor Loadings and reliability of the EQ-SF Items across times.

|  | **Subscales** | **Items** | **T<sub>1</sub>** | **T<sub>2</sub>** | **$\alpha_1$** | **$\alpha_2$** |
| --- | --- | --- | --- | --- | --- | --- |
| PE | Social positive | 11. It is easier for me to socialize | .84 | .82 | .90 | .89 |
|  |  | 19. I am friendlier | .87 | .86 |  |  |
|  |  | 22. I feel more social | .91 | .87 |  |  |
|  | Full positive | 2. I enjoy the buzz | .58 | .57 | .78 | .78 |
|  |  | 17. It is fun | .79 | .74 |  |  |
|  |  | 23. I feel good | .84 | .85 |  |  |
|  | Sex positive | 7. I become more sexually active | .75 | .78 | .86 | .86 |
|  |  | 13. I am more sexually responsive | .92 | .86 |  |  |
|  |  | 18. I am more sexually assertive | .81 | .82 |  |  |
|  | Tension reduction | 5. It take away my negative moods and feelings | .68 | .70 | .69 | .80 |
|  |  | 9. I feel less stressed | .65 | .73 |  |  |
|  |  | 15. I am able to take my mind off my problems | .64 | .83 |  |  |
| NE | Social negative | 1. I become aggressive | .75 | .70 | .68 | .68 |
|  |  | 6. I get into a fights | .67 | .71 |  |  |
|  |  | 14. I get mean | .57 | .55 |  |  |
|  | Emotional negative | 3. I feel ashamed of myself | .74 | .67 | .75 | .75 |
|  |  | 8. I feel guilty | .78 | .81 |  |  |
|  |  | 16. I feel sad or depressed | .62 | .63 |  |  |
|  | Physical negative | 4. I feel sick | .55 | .53 | .76 | .71 |
|  |  | 10. I get a hangover | .82 | .68 |  |  |
|  |  | 20. I get a headache | .80 | .80 |  |  |
|  | Cognitive negative | 12. I become clumsy or uncoordinated | .57 | .50 | .74 | .72 |
|  |  | 21. I can't concentrate | .75 | .77 |  |  |
|  |  | 24. I have problems with memory and concentration | .81 | .76 |  |  |

*Note.* PE = Positive expectancies; NE = negative expectancies; T<sub>1</sub> = Time 1; T<sub>2</sub> = Time 2. $\alpha$ = Cronbach's alpha

### Longitudinal Measurement Invariance of the EQ-SF

The results of the longitudinal measurement invariance analysis across the two assessment times are shown in Table 3. Configural invariance showed acceptable to optimal fit indices. Metric and scalar invariance across times were also found, as changes in fit indices were lower than the specified cut-off criteria. These findings suggest that increasing constraints (i.e., in the item factor loadings and intercepts) across the two time points (T1 and T2) did not significantly worsen the model fit and that LMI was held.

**Table 3.** Model Fit Statistics for the Different Levels of Measurement Invariance of the EQ-SF across times.

| Model | CFI | TLI | RMSEA | SRMR | ΔCFI | ΔTLI | ΔRMSEA | $\Delta$ SRMR |
| --- | --- | --- | --- | --- | --- | --- | --- | --- |
| Configural invariance | 0.920 | 0.904 | 0.045 | 0.057 |  |  |  |  |
| Metric invariance | 0.920 | 0.906 | 0.045 | 0.058 | 0 | -0.002 | 0 | -0.001 |
| Scalar invariance | 0.916 | 0.903 | 0.046 | 0.059 | 0.004 | 0.003 | -0.001 | -0.001 |

*Note.* *CFI* = Comparative Fit Index; *TLI* = Tucker-Lewis Index; *RMSEA* = Root Mean-Square Error of Approximation; *SRMR* = Standardized Root Mean-Square Residual.

### Criterion-related validity

Bivariate correlations are presented in Table 4. Small- and medium-sized significant positive correlations were found between alcohol expectancies at baseline, alcohol-related outcomes, and drinking motives at follow-up. The strongest correlations were observed between positive expectancies and social and enhancement motives.

**Table 4.**
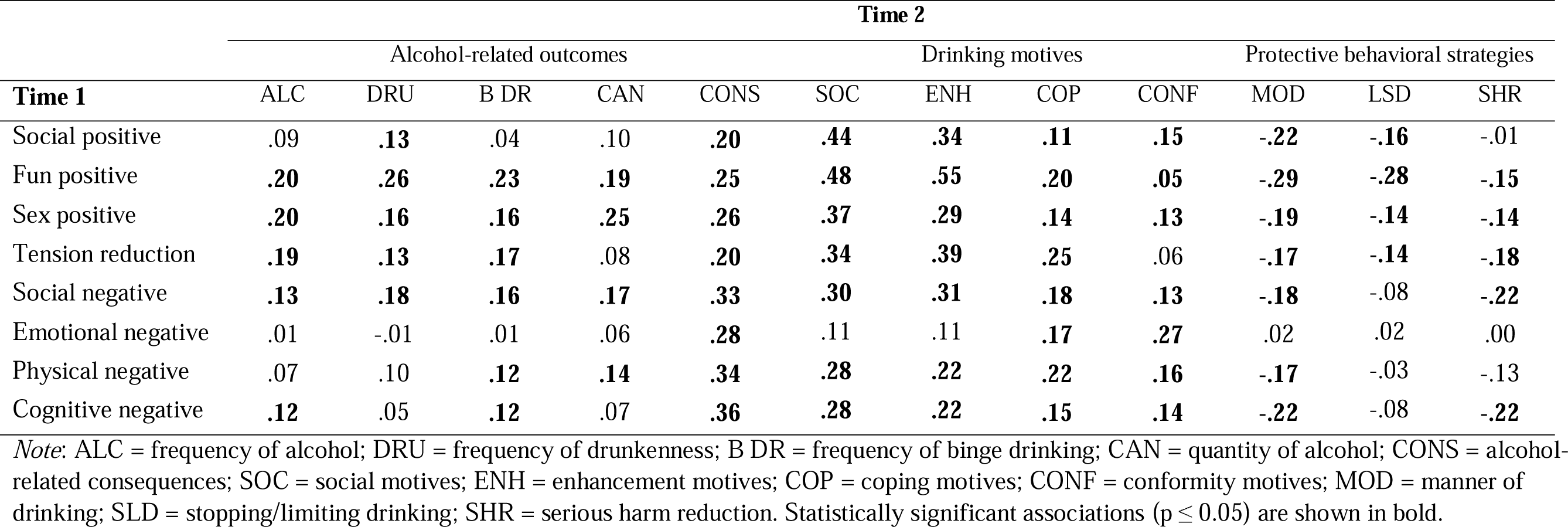
Bivariate correlations between EQ-SF subscale at time 1 with and alcohol-related outcomes indicators (frequency of alcohol use, drunkenness, and binge drinking, quantity of alcohol, and alcohol-related negative consequences), dimensions of drinking motives, and use of PBS at time 2.

Further, most alcohol expectancies at T1 (except emotional negative) showed significant and negative correlations with use of PBS (T2).

## Discussion

The present study examined the reliability, validity, and longitudinal measurement invariance of the Spanish Alcohol Expectancy Questionnaire Short Form (EQ-SF) (25) in a sample of young adults binge drinkers. The results of the CFA revealed that, although both models evaluated (i.e., the eight-factor correlated model and the eight-factor hierarchical model grouped into two general factors of positive and negative expectancies) showed acceptable fit indices, the eight-factor correlated model showed a significantly better fit than the two-factor second-order model. This finding contrasts with previous studies conducted with college students (14), adolescents (24), and adults (25), which have reported a better fit for the hierarchical two-factor (positive and negative) second-order model. Previous research (e.g., (51)) has demonstrated that the relationships between specific dimensions of alcohol expectancies (e.g., fun positive) and drinking behavior may differ between various subpopulations, which could explain the variation in model fit results observed in our study compared to previous findings. The information derived from the subscales can serve to identify young adults with specific beliefs about the effects of alcohol, which may be targeted in interventions. Our findings support the use of specific sub-scales (e.g., (52), for social positive) either alongside or instead of the total score (positive or negative, e.g., (53)) in community-based samples of young adults who engage in binge drinking patterns.

A novel and relevant finding of the present study is that we found evidence to support the temporal invariance of the Spanish version of the EQ-SF. Longitudinal measurement invariance is a central issue in psychological assessment, particularly in the context of repeated assessments (e.g., interventions aimed at reducing alcohol use by reducing positive alcohol expectancies). Unless temporal invariance is met, the meaningful comparison of correlations and mean scores across different time points is unattainable (54). Notably, our research findings establish the presence of configural, metric, and scalar invariance in the multidimensional structure of the EQ-SF across two measurement waves (spaced two months apart). These results suggest the utility of the scale as a reliable instrument for assessing alcohol expectancies in a similar way over time in a high-risk Spanish-speaking population. Given these findings, it is possible to compare mean scores across different time points, and any observed differences can be accurately interpreted as genuine changes in alcohol expectancies.

Additionally, we examined the reliability of the EQ-SF scores and explored evidence of concurrent validity. Alpha coefficients indicated acceptable values (.68 ≤ α ≤ .90) of internal consistency for the EQ-SF dimensions, which aligns with previous findings (e.g., (25); 77 ≤ α ≤ .90) suggesting that the EQ-SF is a reliable measure for assessing alcohol expectancies in young adults who engage in binge drinking patterns. The relationship between EQ-SF scores and other variables, including alcohol-related outcomes (e.g., frequency of alcohol use, frequency of drunkenness, frequency of binge drinking, quantity of alcohol consumed, and alcohol-related consequences), drinking motives, and PBS use, provide evidence of concurrent validity. Specifically, and in accordance with findings from previous studies (e.g., (16)), we observed positive associations between scores on all four factors measuring positive expectancies and indicators of alcohol consumption, such as frequency of alcohol use, frequency of binge drinking, and alcohol-related consequences. Furthermore, our study revealed a positive relationship between the four scales of positive expectancies and a higher frequency of drunkenness. Additionally, the positive correlation between alcohol expectancies and drinking motives is consistent with the postulates of the Motivational Model of Alcohol Consumption (55, 56).

Finally, and in line with previous research conducted with college students (for a review, see 18), individuals with more positive alcohol expectancies were less likely to engage in PBS use, increasing the likelihood of experiencing alcohol-related problems. However, our findings do not support the negative relationship shown in previous studies (e.g., (24)) between negative alcohol expectancies (for most of its dimensions) and drinking behaviors. From a psychometric perspective, the absence of such a relationship may be explained by the fact that a substantial body of research (for a systematic review, see 57) has failed to establish a clear and consistent relationship between negative expectancies and outcomes related to alcohol consumption.

## Limitations and Future Directions

The results of our study should be interpreted in light of certain limitations. One limitation stems from the use of a non-probabilistic sample, given the absence of a sampling frame of young adults who binge drink within community settings. This limitation restricts the generalizability of our findings to the wider population of Spanish young adults. Nonetheless, it is worth noting that the gender distribution of our sample, with 47% females, closely aligns with that of the Spanish young adult population aged 18-25 years (reported as 48.8% females by the National Statistics Institute, 58). Moreover, we recruited both university and non-university students, enhancing the diversity of our sample.

Additionally, although our study utilized a relatively short time frame (last two months) to collect follow-up data, the validity of our results may have been affected by certain self-report biases, particularly recall bias. To mitigate these potential biases and further test the validity of our findings, future research could explore the use of Ecological Momentary Assessments (e.g., (59), (60)).

## Conclusions

The results of this study provide favorable psychometric evidence regarding the reliability and validity of the Spanish EQ-SF in a community sample of Spanish young adult binge drinkers. Furthermore, our findings suggest that the brief version of the EQ- SF is a valuable tool for examining changes in expectancies over time that can be targeted in interventions (e.g., (61)) aimed at young adults who binge drink.

## Data Availability

All data produced in the present study are available upon reasonable request to the authors

